# Serendipitous detection of *Anopheles stephensi* in Kisumu, Kenya in June 2022

**DOI:** 10.1101/2023.05.02.23289394

**Authors:** Bryson Alberto Ndenga, Sammy Wambua, Kevin Omondi Owuor, Rodney Omukuti, Salome Chemutai, Daniel Arabu, Irene Miringu, Carren Bosire, Kavinya Mwendwa, Christabel Achieng Winter, Francis Maluki Mutuku, Donal Bisanzio, Angelle Desiree LaBeaud, Keli Nicole Gerken

## Abstract

In June 2022, a pool of five mosquitoes that were morphologically classified as *Anopheles gambiae* and caught in Kisumu (Kenya) were tested for blood-meal analysis. Of the 19.6% (11/56) amplicon sequence variants assigned to mosquito species using basic local alignment search tool (BLAST), one had 15 hits matching *Anopheles stephensi*.

## Research letter

*Anopheles stephensi* is a native urban malaria vector in south Asia and the Arabian Peninsula. It is capable of transmitting both *Plasmodium falciparum* and *P. vivax* and is projected to expand further as urbanization increases, especially in Africa (*1*). It was first reported in Africa in Djibouti in 2012 (*2*) then in Ethiopia in 2019 (*3*), Sudan in 2018 (*4*), Somali-land in 2020 (*5*), Nigeria in 2020 (*6*) and for the first time in Marsabit County in northern Kenya in December 2022 (*7*). This demonstrates a clear and rapid expansion of its geographical range within the continent.

During a One Health study on livestock and vector dynamics at urban slaughterhouses, we carried out mosquito collections using ovitraps, larval/pupae sampling, Prokopack aspirator and Biogents (BG)-trap baited with CO_2_. This study was titled “Urban Rift Valley Fever Virus as a New Ecological Niche: Continuous Introduction from Animal Products” and performed at the Mamboleo Slaughterhouse (0° 3’ 25.4’’ N, 34° 47’ 10.9’’ E, Figure) in Kisumu City, Kenya, and was approved by Stanford University (61386), Kenya Medical Research Institute Scientific and Ethics Review Unit (KEMRI/SERU/CGHR/03-07-390/4293) and obtained research license from the National Commission for Science, Technology & Innovation (NACOSTI/P/21/13557).

**Figure 1.**
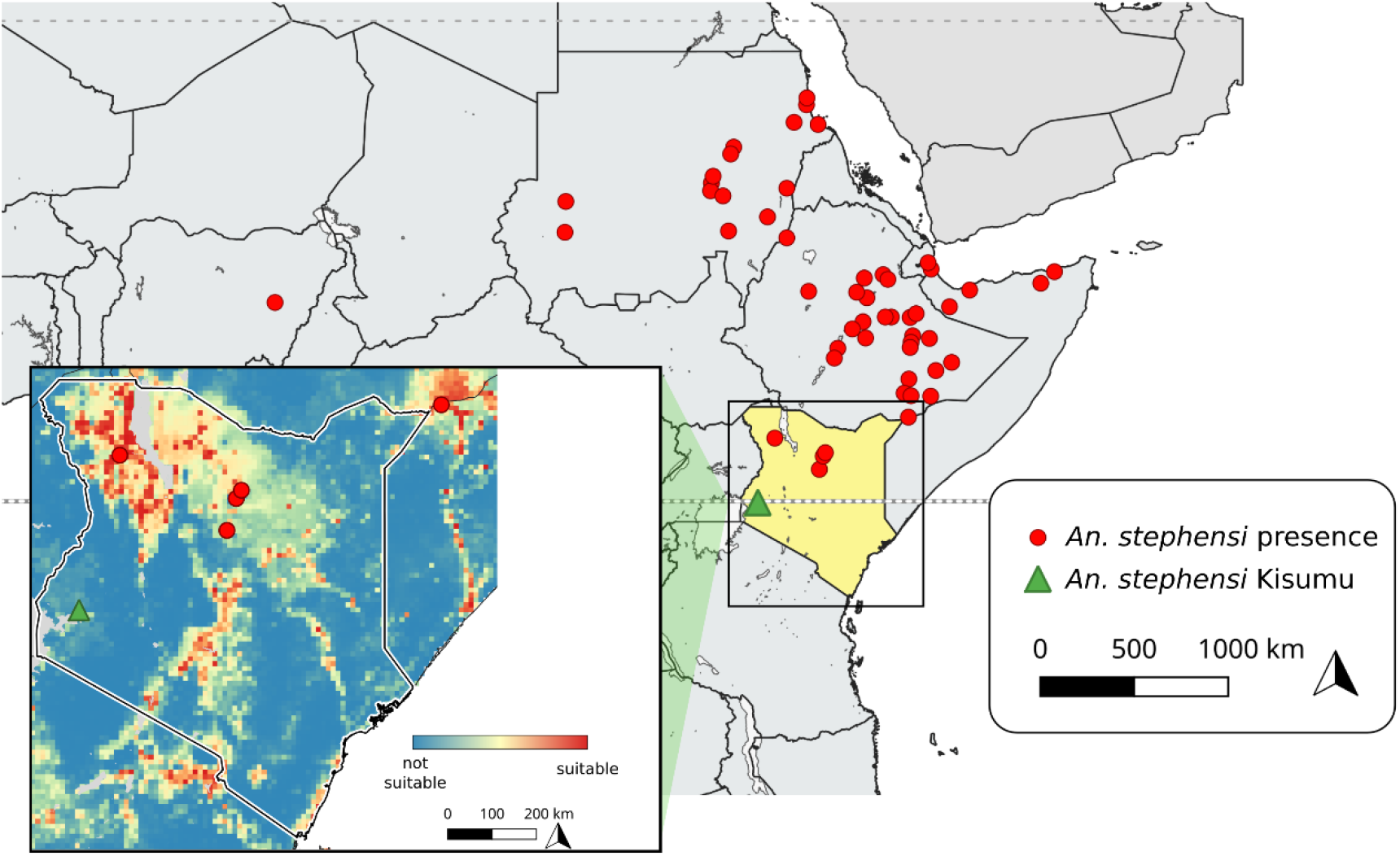
Map showing the location in Kisumu, Kenya where *Anopheles stephensi* (green triangle) was collected in June 2022 and other sites in Africa where it has been reported (Source: *Anopheles stephensi* reports (red points): https://apps.who.int/malaria/maps/threats/). Suitability map created using data obtained from Sinka et al. 2020^1^.

Adult mosquitoes resting indoors and outdoors were sampled using a Prokopack aspirator from May 16^th^ 2022 to July 1^st^ 2022. Sampling lasted one hour and was conducted between 08:45 HRS and 01:30 HRS. Subsequent samplings were completed biweekly. Collected mosquitoes were killed using a pyrethrum aerosol spray before morphological identification in a field station and recorded on a data form. Female mosquitoes were further sorted according to their blood-feeding stages as unfed, blood-fed, half-gravid or gravid. Mosquitoes were preserved in silica gel self-indicating 6-20mesh (Blue) - 500gm (Loba Chemie Pvt. Ltd, Jehangir Villa, 107, Wodehouse Road, Colaba, Mumbai 400 005, India) with the intention for further blood meal testing.

A total of 2,891 mosquitoes were collected: 66.2% (1,782) indoors and 33.8% (765) outdoors; 50.7% (1,465) males and 49.3% (1,426) females. Species identified morphologically were: 0.3% (9) *Aedes aegypti*; 6.3% (182) *Anopheles coustani*; 0.3% (8) *An. funestus*; 10.9% (315) *An. gambiae*, and 82.2% (2,377) *Culex* spp. Over a half, 61.6% (194/315), of the *An. gambiae* were collected outdoors and 38.4% (121) indoors; 46.3% (146) were males; 19.0% (60) un-blood-fed; 25.1% (79) blood-fed; 6.0% (19) half-gravid and 3.5% (11) gravid. A sample of the blood-fed mosquitoes were transported on dry ice to Pwani University Biosciences Research Centre (PUBReC) in Kilifi, Kenya, for determination of blood meal sources.

Genomic DNA was extracted from the mosquito abdomens using the TIANamp Genomic DNA Kit (Tiangen, Beijing, China). Quality and quantity of DNA was analyzed by NanoDrop 2000C spectrophotometer (Thermo Scientific Inc, USA) and verified on 1% agarose gel electrophoresis. To identify blood meal sources, we PCR-amplified ∼300 base pair (bp) of the cytochrome b barcode (*8*) which was high-throughput-sequenced, at Macrogen Inc., Seoul Korea, using the Illumina 300 × 2 bp platform (Illumina, USA). The amplicon sequence data were analyzed with the DADA2 (version 1.21.0) bioinformatics pipeline (Callahan et al., 2016) implemented in the R programming language (version 4.1.1). After quality preprocessing, high quality reads were denoised and clustered into unique sequences i.e., amplicon sequence variants (ASVs). We assigned taxonomy to the ASVs using two approaches. We assigned taxonomy, first, by exact ASV matching against DADA2-trained MIDORI2 reference database (version GB254) which includes eukaryotic mitochondrial sequences (*9*). ASVs that were not assigned by this approach were compared against the NCBI database using BLAST (*10*). Raw sequences are available in SRA database of NCBI under project accession number PRJNA966766.

In all, 248 ASVs were inferred, 192 of which were classified to 14 vertebrate species (Mammalia and Aves) by exact matching against MIDORI2 reference database. Other than vertebrates, 19.6% (11/56) of the ASVs assigned using BLAST matched to mosquito species one of which was *An. stephensi* with a total of 15 hits (89-93% identity, 67-85% coverage, E-value < 1e-46 (Table). The reads matching *An. stephensi* were tracked to a pool of five (5) mosquitoes that had been morphologically classified as *An. gambiae* in the field. These mosquitoes were collected from Mamboleo Slaughterhouse in Kisumu City, Kenya in June 2022.

**Table 1.** Summary of coverage and identity values for each of the top ten (10) hits in the BLAST search matching *Anopheles stephensi*.

| Description | Maximum score | Query coverage | E value | Identity | Accession |
| --- | --- | --- | --- | --- | --- |
| *Anopheles stephensi* strain Indian chromosome 2L | 326 | 84% | 2.00E-84 | 92.51% | CP032300.1 |
| *Anopheles stephensi* strain Indian chromosome 3L | 320 | 85% | 8.00E-83 | 92.07% | CP032302.1 |
| *Anopheles stephensi* strain Indian chromosome 3R | 320 | 82% | 8.00E-83 | 92.07% | CP032301.1 |
| *Anopheles stephensi* strain Indian chromosome 2R | 315 | 81% | 4.00E-81 | 91.63% | CP032299.1 |
| *Anopheles stephensi* strain SDA-500 chromosome 3R | 315 | 83% | 4.00E-81 | 91.63% | CP032234.1 |
| *Anopheles stephensi* strain SDA-500 chromosome 2R | 309 | 81% | 2.00E-79 | 91.19% | CP032232.1 |
| *Anopheles stephensi* strain SDA-500 chromosome 2L | 303 | 84% | 9.00E-78 | 90.75% | CP032233.1 |
| *Anopheles stephensi* strain SDA-500 chromosome 3L | 298 | 88% | 4.00E-76 | 90.31% | CP032235.1 |
| *Anopheles ziemanni* genome assembly, chromosome: 2 | 291 | 83% | 7.00E-74 | 89.52% | OX030919.2 |

Our findings report the first detection of *An. stephensi* south of the Equator. Furthermore, this detection represents the presence of an invasive vector in an urban area that is malaria endemic. Our study suggests that the detections of *An. stephensi* within Africa (*1-5, 7*) could be dramatically underreported. The initial false identification of this vector also indicates that to fully capture this vector’s boundary, molecular methods and confirmatory testing may be required. The presence of a malaria vector adapted to breeding in open water containers prevalent in urban areas is particularly concerning. *Anopheles stephensi* can effectively transmit *P. falciparum* and *P. vivax* and entry into malaria endemic zones could shift malaria transmission dynamics considerably.

## Data Availability

All data produced in the present study are available upon reasonable request to the authors

## Acknowledgements

We thank Charles Otieno Adipo for sampling the vectors, the management of Mamboleo Slaughterhouse for allowing us to sample mosquitoes in the premises and our colleagues in KEMRI and Stanford University for their support to approve this research study. This study was funded by the Stanford University Center for Innovations in Global Health Seed Grant 2021 and the Robert E Shope ASTMH (American Society of Tropical Medicine and Hygiene) to Dr. Keli Nicole Gerken.

## About the Author

Bryson Alberto Ndenga, PhD is a Senior Research Scientist in Kenya Medical Research Institute, Centre for Global Health Research. His primary interest is in the biology, ecology and control of mosquitoes that transmit malaria, dengue, chikungunya and Rift Valley fever.

